# Language disorder in progressive supranuclear palsy and corticobasal syndrome: neural correlates and detection by the MLSE screening tool

**DOI:** 10.1101/2021.03.04.21252852

**Authors:** Katie A. Peterson, P. Simon Jones, Nikil Patel, Kamen A. Tsvetanov, Ruth Ingram, Stefano F. Cappa, Matthew A. Lambon Ralph, Karalyn Patterson, Peter Garrard, James B. Rowe

**Author notes:** **Correspondence:** Professor James Rowe, Department of Clinical Neurosciences, University of Cambridge, Herchel Smith Building for Brain and Mind Sciences, Forvie Site, Robinson Way, Cambridge CB2 0SZ, UK;.

## Abstract

**Background:** Progressive supranuclear palsy (PSP) and corticobasal syndrome (CBS) affect speech and language as well as motor functions. Clinical and neuropathological data indicate a close relationship between these two disorders and the non-fluent variant of primary progressive aphasia (nfvPPA). We use the recently developed Mini Linguistic State Examination tool (MLSE) to study speech and language disorders in patients with PSP, CBS, and nfvPPA, in combination with structural magnetic resonance imaging (MRI).

**Methods:** Fifty-one patients (PSP N = 13, CBS N = 19, nfvPPA N = 19) and 30 age-matched controls completed the MLSE, the short form of the Boston Diagnostic Aphasia Examination (BDAE), and the Addenbrooke’s Cognitive Examination III. Thirty-eight patients and all controls underwent structural MRI at 3 Tesla, with T1 and T2-weighted images processed by surface-based and subcortical segmentation within FreeSurfer 6.0.0 to extract cortical thickness and subcortical volumes. Morphometric differences were compared between groups and correlated with severity of speech and language impairment.

**Results:** CBS and PSP patients showed impaired MLSE performance, compared to controls, with a similar language profile to nfvPPA, albeit less severe. All patient groups showed reduced cortical thickness in bilateral frontal regions and striatal volume. PSP and nfvPPA patients also showed reduced superior temporal cortical thickness, with additional thalamic and amygdalo-hippocampal volume reductions in nfvPPA. Multivariate analysis of brain-wide cortical thickness and subcortical volumes with MLSE domain scores revealed associations between performance on multiple speech and language domains with atrophy of left-lateralised fronto-temporal cortex, amygdala, hippocampus, putamen and caudate.

**Conclusions:** The effect of PSP and CBS on speech and language overlaps with nfvPPA. These three disorders cause a common anatomical pattern of atrophy in the left frontotemporal language network and striatum. The MLSE is a short clinical screening tool that can identify the language disorder of PSP and CBS, facilitating clinical management and patient access to future clinical trials.

## Introduction

Progressive supranuclear palsy (PSP) and corticobasal degeneration are primary degenerative tauopathies affecting movement and cognition (Litvan et al., 1996; Armstrong et al., 2013; Burrell et al., 2014; Höglinger et al., 2017). Speech and language deficits are common in both disorders, but their recognition has been hampered by the lack of a brief but sensitive clinical assessment tool. In this paper, we use the Mini Linguistic State Examination (MLSE (Patel et al., 2020)) to investigate the range of speech and language deficits in PSP and the corticobasal syndrome (CBS), and their neural correlates in structural magnetic resonance imaging (MRI).

The classical phenotype of PSP, Richardson’s syndrome, is characterised by vertical supranuclear gaze palsy, axial rigidity, and postural instability, with cognitive impairment (Steele et al., 1964; Litvan et al., 1996). Richardson’s syndrome is highly suggestive of PSP pathology, however, other common phenotypes have been described, including presentation with speech and language deficits (PSP-SL) (Respondek and Höglinger, 2016; Höglinger et al., 2017). The clinical syndrome of CBS is characterised by the combination of motor deficits (progressive asymmetrical akinetic rigidity, dystonia, tremor, myoclonus) and cortical features (alien limb, apraxia, cortical sensory change) (Riley and Lang, 1988; Boeve et al., 2003), with heterogeneity in the clinical presentations and underlying pathology (Armstrong et al., 2013; Alexander et al., 2014). CBS is commonly accompanied by impaired speech and language (Burrell et al., 2014; Peterson et al., 2019). Indeed, a non-fluent agrammatic presentation of CBS (CBS-NAV) is recognised in consensus clinical diagnostic criteria (Armstrong et al., 2013). Speech and language deficits also develop commonly in PSP and CBS after motor presentations (Catricalà et al., 2019; Dodich et al., 2019; Peterson et al., 2019).

The speech and language changes of PSP and CBS have much in common with the non-fluent variant of primary progressive aphasia (nfvPPA). Indeed, there is considerable overlap in the criteria for diagnosing nfvPPA, PSP-SL, and CBS-NAV (Peterson et al., 2019). There is agrammatism, anomia, circumlocution, and impaired syntactic comprehension in the context of preserved single-word comprehension and object knowledge in patients with PSP-SL and CBS-NAV (Armstrong et al., 2013; Höglinger et al., 2017; Dodich et al., 2019), and subtle deficits in verbal production and sentence comprehension in PSP/CBS (Dodich et al., 2019). However, few studies have directly compared speech and language in PSP/CBS with nfvPPA. Burrell et al. (Burrell et al., 2018) found aphasic deficits on formal testing in PSP patients that were comparable in frequency and severity to those of an nfvPPA group. However, the PSP group were recruited mainly from a cognitive disorders clinic and may have overrepresented cognitive phenotypic presentations.

Although the speech and language changes of PSP and CBS have similarities to nfvPPA, it does not necessitate a common aetiology. However, there are also overlapping neuropathological features including neuronal, oligodendroglial, and astrocytic inclusions that are immunoreactive for tau with 4 microtubule binding repeats (4R) (Grossman, 2010; Dickson et al., 2011; Spinelli et al., 2017). PSP and CBS clinical signs often follow the presentation of nfvPPA (Kertesz and McMonagle, 2010; Santos-Santos et al., 2016; Cerami et al., 2017; Gazzina et al., 2019), or primary progressive apraxia of speech (Josephs et al., 2014). The clinical and pathological overlap of PSP, CBS, and nfvPPA underlies the concept of a continuous spectrum of 4R-Tauopathy disorders (Kertesz et al., 2005; Dickson et al., 2011; Murley et al., 2020) that extends to the functional anatomy of their cognitive deficits.

Here, we test the hypothesis that the three disorders have a common associated structural impairment in relation to their common effects on speech and language. The functional anatomy of language impairment in PSP and CBS has been identified by fluorodeoxyglucose positron emission tomography (Dodich et al., 2019). Patients with language presentations of these two conditions have hypometabolism in areas typical of nfvPPA (left fronto-insular and superior medial frontal cortex), whereas patients without language impairment showed predominant right hemispheric involvement. At the group level, the disorders differ: PSP is associated with atrophy and hypometabolism of midbrain, striatal, and frontal regions, bilaterally (Kaat et al., 2011; Niccolini and Politis, 2016; Murley et al., 2020); CBS is associated with asymmetric hypometabolism and atrophy of fronto-parietal cortex and basal ganglia (Niccolini and Politis, 2016; Murley et al., 2020); and nfvPPA is associated with atrophy and hypometabolism of the left frontal perisylvian region, anterior insula and frontal operculum (Nestor et al., 2003; Gorno-Tempini et al., 2004; Preiß et al., 2019).

The MLSE was recently developed as a screening tool to identify and categorise speech and language deficits in neurological disorders (Patel et al., 2020). We therefore used the MLSE to compare PSP, CBS and nfvPPA. The speech and language symptoms of PSP and CBS have been difficult to assess and characterise in non-specialist settings because of the lack of a validated brief language screening test. The MLSE is accurate for the assessment of primary progressive aphasias (Patel et al., 2020) but its performance in other neurological disorders is yet to be assessed. We included patients with a range of PSP and CBS phenotypic presentations to reflect the range of cases presenting to cognitive and movement disorders clinics, noting that speech and language impairment occurs in PSP/CBS patients even in those who do not meet criteria for PSP-SL or CBS-NAV (Dodich et al., 2019).

This study had two key aims: (1) to use the MLSE to evaluate and compare linguistic impairment in PSP, CBS and nfvPPA; and (2) to investigate brain structural correlates of speech and language deficits in PSP, CBS and nfvPPA. We predicted that the language profile in PSP and CBS would resemble that seen in nfvPPA. We also predicted that performance on the MLSE would be associated with cortical atrophy in a left-lateralised language network, specifically the inferior frontal cortex, associated with agrammatic and apraxic speech (Gorno-Tempini et al., 2004).

## Materials and Methods

### Ethics

Ethical approval was obtained from the London-Chelsea Research Ethics Committee (REC reference: 16/LO/1735). The study was sponsored by St George’s, University of London. All participants provided written informed consent.

### Participants

Fifty-one patients (PSP N = 13, CBS N = 19, nfvPPA N = 19) were recruited through specialist cognitive neurology clinics at Addenbrooke’s Hospital, Cambridge (N = 33), St George’s Hospital, London (N = 13), and Manchester Royal Infirmary and its associated clinical providers (N = 5). Patients were included if they had a clinical diagnosis of PSP based on the 2017 Movement Disorder Society criteria (Höglinger et al., 2017), CBS based on the 2013 Armstrong et al. criteria (Armstrong et al., 2013), or nfvPPA based on the 2011 Gorno-Tempini criteria (Gorno-Tempini et al., 2011). Nine of the PSP patients had probable PSP-Richardson’s syndrome; the other four included one each of probable PSP-progressive gait freezing (PSP-PGF), probable PSP-SL, possible PSP-ocular motor dysfunction (PSP-OM), and possible PSP. Eight CBS patients had probable CBS; the others had probable CBS-NAV (N = 6), possible CBS-NAV (1), probable CBS presenting as PSP syndrome (CBS-PSP) (1), and possible CBS (3). One nfvPPA patient declared a native language other than English but had been highly fluent in English since childhood and predominately used English in day-to-day life. Patients were excluded if they had advanced dementia and were unable to understand the purpose of the study or follow task instructions.

Thirty healthy controls were recruited. Inclusion criteria for the healthy control group were: the absence of a diagnosis of any pathological process causing a cognitive disorder and/or subjectively reported cognitive decline; age between 40 and 75 years; English as a first language; willing to participate in a study investigating language and dementia. Healthy controls were recruited through the National Institute for Health Research “Join Dementia Research” register (www.joindementiaresearch.nihr.ac.uk) in Cambridge and London, patients’ relatives, and via local advertisement.

### Cognitive and language assessment

Participants completed the Addenbrooke’s Cognitive Examination – III (ACE-III) (Hsieh et al., 2013), the short form of the Boston Diagnostic Aphasia Examination (BDAE) (Goodglass et al., 2001), and the Mini Linguistic State Examination (MLSE) (Patel et al., 2020). The short form of the BDAE, designed to assess language changes after focal brain damage (mainly stroke), was used to augment the MLSE assessment of language functions. A composite score was calculated from the following subtests of the short form of the BDAE, selected to overlap with the MLSE subtasks: single word repetition (max = 5), basic word discrimination (max = 16), sentence repetition (max = 2), the Boston Naming Test short form (max = 15), and basic oral word reading (max = 15), giving a maximum BDAE composite score of 53 to compare with the MLSE total score. Assessment sessions were video and/or audio recorded for offline scoring and analysis.

The MLSE is a brief language assessment tool designed for the assessment of progressive aphasia. It contains subtests which span the principal linguistic domains affected by PPA, as used to apply the diagnostic criteria (Gorno-Tempini et al., 2011): confrontation naming, repetition, word and sentence comprehension, semantic association, reading, writing and a connected speech task. Error-based scoring provides *domain scores* corresponding to key linguistic domains (motor speech, knowledge of phonological structure, semantic knowledge, syntactic knowledge, and auditory-verbal working memory) as well as an overall *total score* out of 100, with lower scores indicating poorer performance. The MLSE has shown high inter-rater agreement and diagnostic accuracy for the classification of primary progressive aphasic syndromes (>90% accuracy using random forest classification) (Patel et al., 2020).

### Data management

Study data were collected and managed using the Research Electronic Data Capture tool, a secure, web-based software platform designed to support data capture for research studies, hosted at University of Cambridge and at St George’s, University of London (Harris et al., 2009, 2019).

### Magnetic Resonance Imaging protocol

Thirty-eight (PSP N = 11, CBS N = 14, nfvPPA N = 13) patients and all controls underwent structural MRI at 3 Tesla with a T1-weighted magnetisation-prepared rapid acquisition gradient echo (MPRAGE) and T2-weighted sequences. Twenty-seven patients (PSP N = 9, CBS N = 9, nfvPPA N = 9) and 20 controls were scanned at the Wolfson Brain Imaging Centre, University of Cambridge on a Siemens Prisma 3T MRI (T1 Sagittal iPAT 2 parameters: TR=2000 ms, TE=2.93 ms, TA=306 s, in-plane resolution = 1.1 x 1.1 mm, slice thickness = 1.1 mm, Inversion Time=850 ms, Flip Angle=8°; T2 Sagittal iPAT 2 parameters: TR=3200 ms, TE=401 ms, TA=283 s, in-plane resolution = 1.1 x 1.1 mm, slice thickness = 1.1 mm, Inversion Time=850 ms, Flip Angle=120 °;). Eleven patients (PSP N = 2, CBS N = 5, nfvPPA N = 4) and 10 controls were scanned at the St George’s Hospital Radiology Department on a Philips Achieva 3T MRI (T1 Sagittal SENSE parameters: TR=6600-6900 ms, TE=3.0-3.2 ms, in-plane resolution = 1.1 x 1.1 mm, slice thickness = 1.1 mm, Inversion Time=850 ms, Flip Angle=8 °; T2 Sagittal SENSE parameters: TR=2200 ms, TE=243 ms, in-plane resolution = 1.1 x 1.1 mm, slice thickness = 1.1 mm, Inversion Time=850 ms, Flip Angle=90 °;).

### Analysis

Demographic, cognitive, and subcortical volumetric data were analysed using RStudio and R version 4.0.2. A chi-square test was conducted to investigate differences in sex distribution between groups. Since Levene’s test showed that the variances for years of education and ACE-III score were not equal, Welch’s ANOVA with Games-Howell *post hoc* test was used. Age, symptom duration, the BDAE composite score, MLSE total and MLSE domain scores, and volumes of thalamus, caudate, nucleus accumbens and brainstem were not normally distributed. Therefore, the Kruskal-Wallis rank sum test was used, with *post hoc* pairwise comparisons by Dunn test. Multiple testing correction was conducted using the Benjamini-Hochberg method (Benjamini and Hochberg, 1995). A corrected value of p<.05 was considered significant.

The T1- and T2-weighted images were processed using FreeSurfer 6.0.0 recon-all pipeline. The T2 volume was used to aid definition of the pial surface. All images were reviewed to confirm accurate segmentation. Group differences in cortical thickness between each patient group and the control group were assessed by t-tests.

We examined the relationship between language impairments and imaging metrics using univariate and multivariate statistics. The univariate approach tested the relationship between MLSE total score and cortical thickness with gender and age as covariates of no interest. Group comparisons and correlational analyses for cortical thickness were performed at each vertex subject to clusterwise correction for multiple comparisons using a permutation analysis with 10,000 randomisations and an initial uncorrected height threshold of *p*<.01. Clusters surviving a two-sided corrected cluster threshold of *p*<.05 were deemed significant. To investigate the link between MLSE and subcortical brain regions, left and right subcortical structure volumes from FreeSurfer were combined. Partial correlations were used to investigate associations between volumes of subcortical structures with MLSE total score, covarying estimated total intracranial volume.

To assess the multivariate relationship between MLSE domain scores and brain structures, we adopted a two-level procedure (Tsvetanov et al., 2018, 2019; Passamonti et al., 2019). First, canonical correlation analysis (Hotelling, 1936) identified the linear relationship between the two multivariate datasets, namely structural values (cortical thickness and subcortical volume) and MLSE domain scores, providing pairs of latent variables (Structure-LV and MLSE-LV). Each latent variable is a linear combination of the original variables, optimised to maximise the correlation between each pair. Here, dataset 1 consisted of structural values of cortical thickness and subcortical volume (67 subjects x 83 nodes: 68 from the Desikan-Killiany Atlas and 15 [left and right: thalamus, caudate, putamen, pallidum, hippocampus, amygdala and nucleus accumbens; and the brainstem] from the automatic subcortical segmentation atlas within FreeSurfer (Fischl et al., 2002)), and dataset 2 included MLSE domain scores (67 subjects x 5 domains). Covariates of no interest included scanner site, gender, age, and total intracranial volume. Next, we tested whether the identified relationship between the cortical thickness and subcortical volume profile (Structure-LV) and MLSE-LV was differentially expressed by groups. We performed a second-level analysis using multiple linear regression with robust fitting algorithm as implemented in the Matlab fitlm.m function. Independent variables included subjects’ brain structure scores from first level canonical correlation analysis, group information (patient vs control) and their interaction (Structure-LV x Group). The dependent variable was subjects’ MLSE-LV scores from the first level analysis in the corresponding canonical correlation analysis.

## Results

### Participant demographics

Fifty-one patients (PSP N = 13, CBS N = 19, nfvPPA N = 19) and 30 controls completed the language assessment, as summarised in Table 1. The groups were similar in age and sex. The patient groups showed similar symptom duration (F<1). There was a significant difference across the groups in years of education. *Post hoc* pairwise comparisons indicated that the control group had significantly more years of education than each patient group (all *p*≤.003), but the patient groups did not differ from each other on this variable. There was a significant difference across the groups in ACE-III total score: higher in the control group than each patient group (all *p*≤.001). In addition, the nfvPPA mean ACE-III total score was lower than PSP (*p* = .019).

**Table 1.**
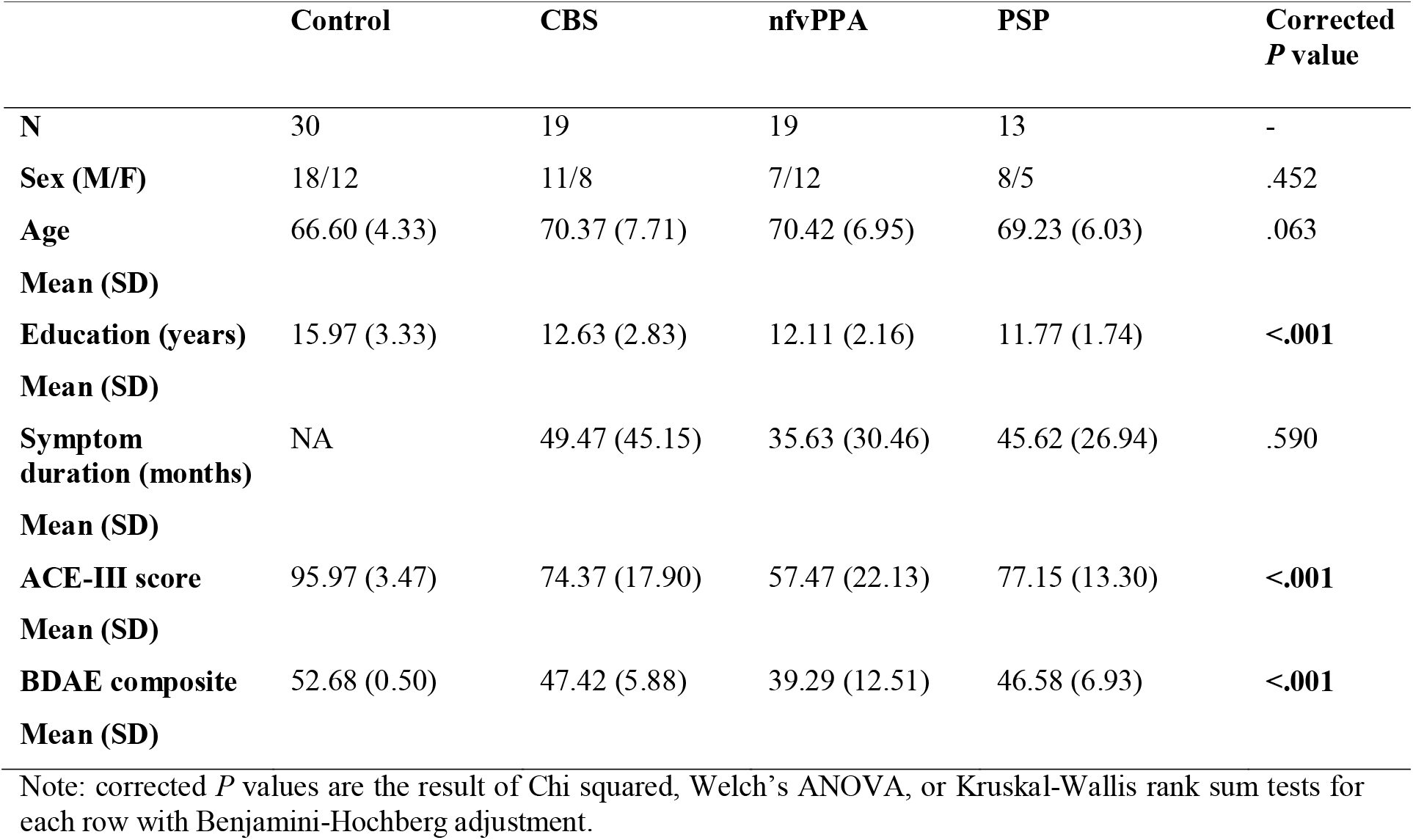
Demographic information for the study cohort

|  | Control | CBS | nfvPPA | PSP | Corrected<br>*P* value |
| --- | --- | --- | --- | --- | --- |
| **N** | 30 | 19 | 19 | 13 | - |
| **Sex (M/F)** | 18/12 | 11/8 | 7/12 | 8/5 | .452 |
| **Age** | 66.60 (4.33) | 70.37 (7.71) | 70.42 (6.95) | 69.23 (6.03) | .063 |
| **Mean (SD)** |  |  |  |  |  |
| **Education (years)** | 15.97 (3.33) | 12.63 (2.83) | 12.11 (2.16) | 11.77 (1.74) | <.001 |
| **Mean (SD)** |  |  |  |  |  |
| **Symptom duration (months)** | NA | 49.47 (45.15) | 35.63 (30.46) | 45.62 (26.94) | .590 |
| **Mean (SD)** |  |  |  |  |  |
| **ACE-III score** | 95.97 (3.47) | 74.37 (17.90) | 57.47 (22.13) | 77.15 (13.30) | <.001 |
| **Mean (SD)** |  |  |  |  |  |
| **BDAE composite** | 52.68 (0.50) | 47.42 (5.88) | 39.29 (12.51) | 46.58 (6.93) | <.001 |
| **Mean (SD)** |  |  |  |  |  |

Note: corrected *P* values are the result of Chi squared, Welch's ANOVA, or Kruskal-Wallis rank sum tests for each row with Benjamini-Hochberg adjustment.

### Language scores

Figure 1 presents group performance for MLSE domains. There were differences in scores between the groups for MLSE total score (χ^2^(3) = 58.52, adjusted *p*<.001), motor speech (χ^2^(3) = 48.90, adjusted *p*<.001), phonological structure (χ^2^(3) = 47.56, adjusted *p*<.001), semantic knowledge (χ^2^(3) = 39.40, adjusted *p*<.001), syntactic knowledge (χ^2^(3) = 46.56, adjusted *p*<.001), and auditory-verbal working memory (χ^2^(3) = 8.16, adjusted *p*=.043). Results of *post hoc* pairwise comparisons using the Dunn test with Benjamini-Hochberg adjustment (Benjamini and Hochberg, 1995) are presented in Figure 1.

**Figure 1.**
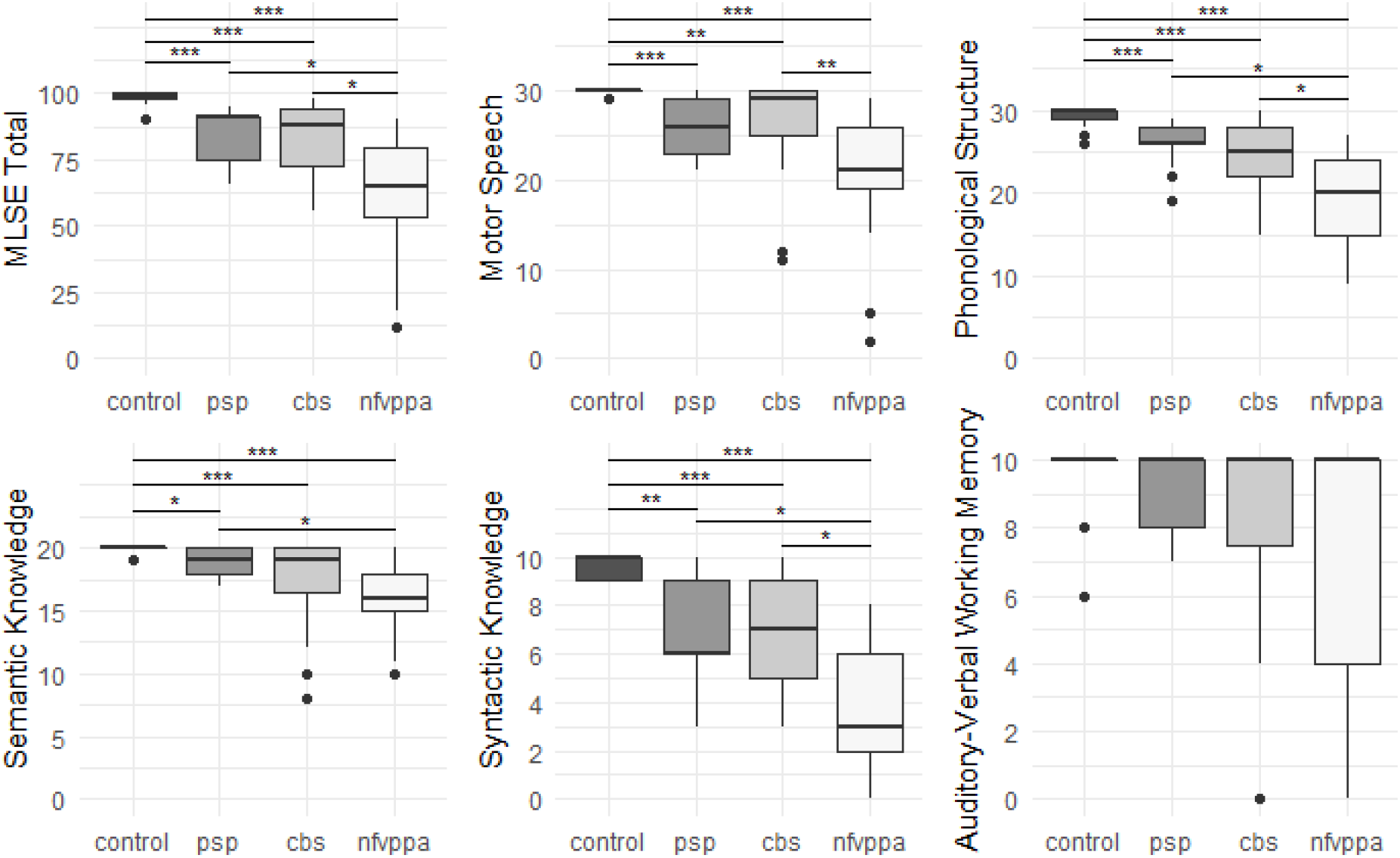
Boxplots of group MLSE total and domain scores. The y-axes for each plot span the min to max scores. Significance markers represent adjusted *p* values from *post hoc* pairwise comparisons using Dunn test with Benjamini-Hochberg adjustment. ‘***’ = adjusted *p* value <.001, ‘**’ = adjusted *p* value <.01, ‘*’ = adjusted *p* value <.05.

Each patient group performed worse than controls for MLSE total score and all MLSE subdomain scores apart from auditory-verbal working memory (after correction for multiple comparisons). The nfvPPA group scored lower than PSP and CBS groups on MLSE total score, with no significant difference between PSP and CBS. The PSP and CBS patients’ MLSE subdomain scores were similar, while nfvPPA patients scored significantly lower than: CBS patients for motor speech, PSP patients for semantic knowledge, and both PSP and CBS patients for phonological structure and syntactic knowledge.

There was a significant difference in BDAE composite score across the groups (χ^2^(3) = 46.63, corrected *p*<.001), with the control group scoring significantly higher than each patient group but with no significant differences between patient groups (see Table 1).

MLSE scores were converted to percent scores to visualise the pattern of linguistic impairment across the groups. As shown in Figure 2, the *pattern* of impairment on MLSE subdomains is comparable across PSP, CBS and nfvPPA but with greater severity in the nfvPPA patients.

**Figure 2.**
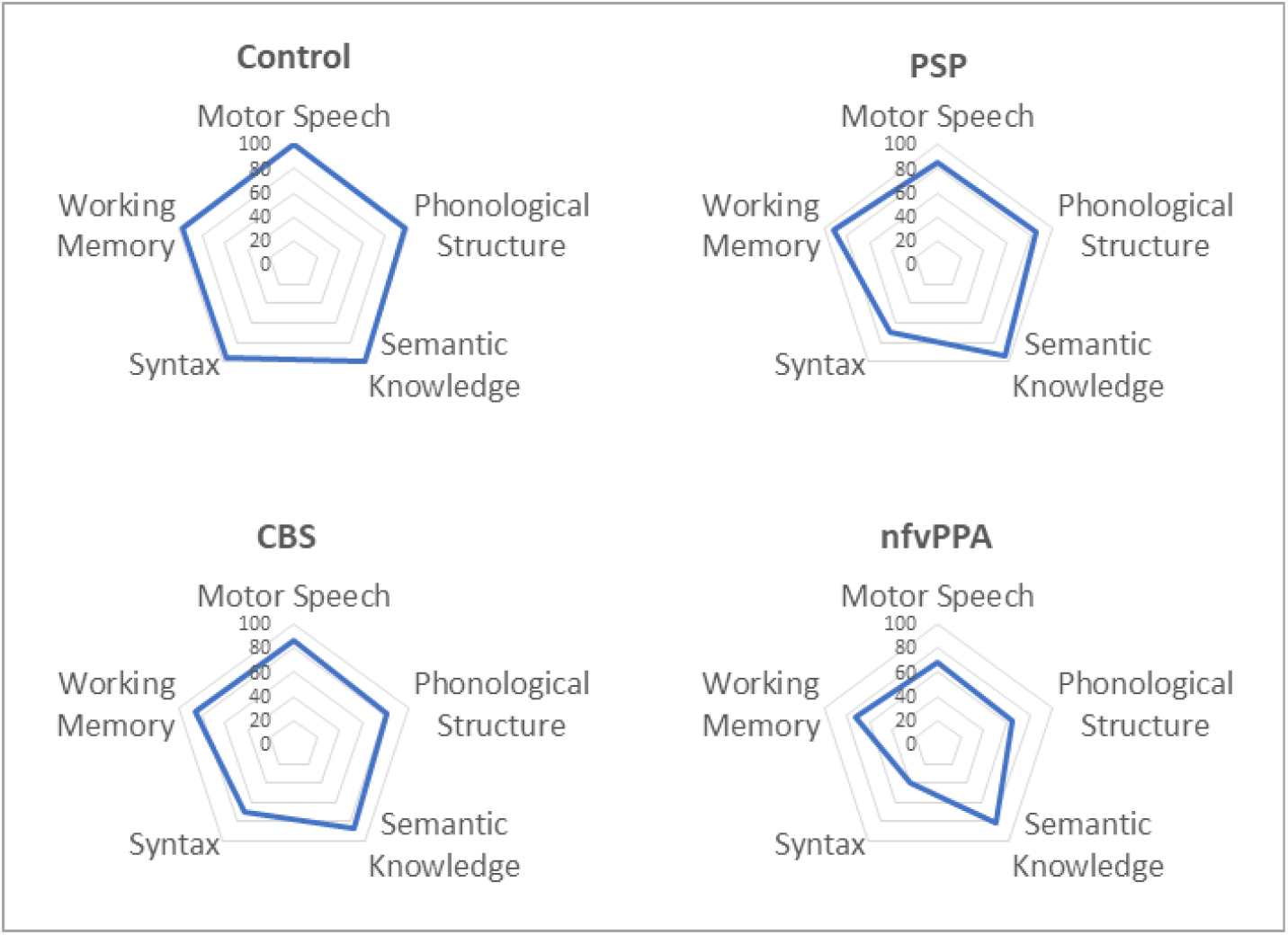
Radar plots showing percentage scores for MLSE subdomains.

### Imaging results

There were differences in cortical thickness between controls and each patient group as shown in Figure 3A. All patient groups showed bilateral cortical thinning in medial and lateral frontal regions, with overlapping regions of cortical thinning in all three patient groups encompassing inferior frontal, middle frontal, superior frontal and precentral gyri, as shown in Figure 3B. The PSP and nfvPPA groups additionally showed cortical thinning in superior temporal regions, bilaterally in PSP and left–sided in nfvPPA. Correlational analysis between the MLSE total score and cortical thickness yielded no significant clusters following clusterwise correction.

**Figure 3.**
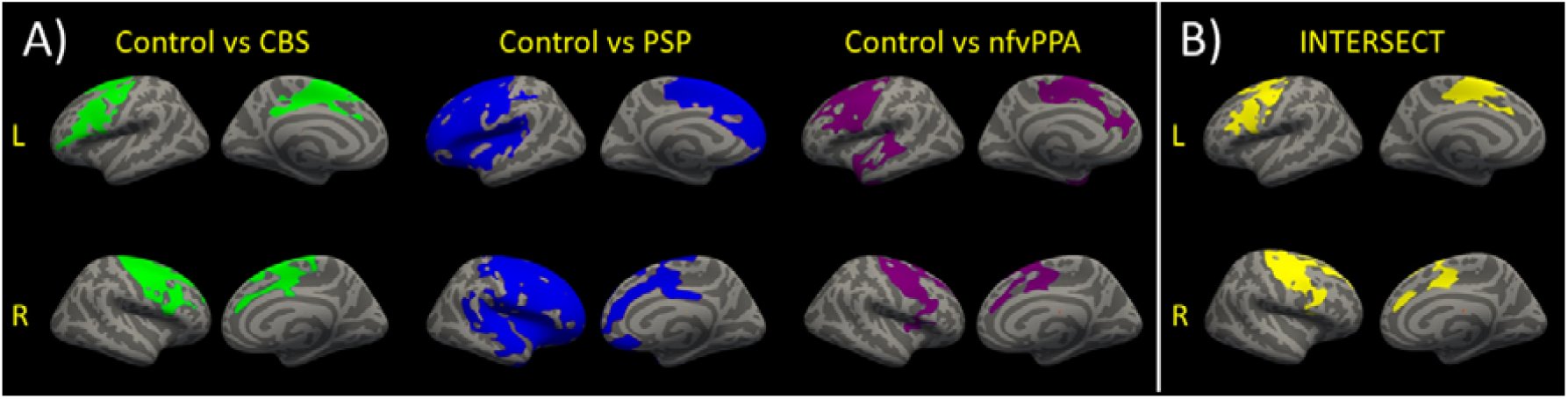
(A) Reduced cortical thickness in each patient group versus the control group. Clusters represent regions of significant differences in cortical thickness between the groups following clusterwise correction. (B) The intersect of the clusters for the three group comparisons of cortical thickness.

There were group differences in subcortical volumes as shown in Table 2. All patient groups showed smaller putamen compared to controls. In addition, PSP patients showed reduced caudate volume compared to controls and reduced pallidum volume compared to all other groups; and nfvPPA patients showed reduced thalamus, caudate, and hippocampus volumes compared to controls and reduced amygdala volume compared to all other groups.

**Table 2.** Subcortical volumes

|  | **Control** | **PSP** | **CBS** | **nfVPPA** |
| --- | --- | --- | --- | --- |
|  | **Mean (SD)** | **Mean (SD)** | **Mean (SD)** | **Mean (SD)** |
| Thalamus | 13.33 (1.47) | 12.73 (3.84) | 12.06 (1.69) | **11.74 (1.51)\*** |
| Caudate | 6.85 (1.08) | **6.01 (1.27)\*** | 6.39 (1.15) | **5.66 (0.82)\*** |
| Putamen | 8.83 (1.05) | **6.87 (1.11)\*** | **7.85 (1.42)\*** | **7.21 (1.00)\*** |
| Pallidum | 3.79 (0.43) | **2.86 (0.38)\*<sup>a, b</sup>** | 3.70 (0.51) | 3.53 (0.54) |
| Hippocampus | 8.06 (0.85) | 7.50 (0.86) | 7.40 (1.10) | **7.11 (1.02)\*** |
| Amygdala | 3.27 (0.45) | 3.14 (0.38) | 2.99 (0.46) | **2.50 (0.47)\*<sup>a, c</sup>** |
| Nucleus<br>Accumbens | 0.96 (0.13) | 0.90 (0.35) | 0.86 (0.18) | 0.82 (0.17) |
| Brainstem | 20.35 (2.17) | 19.99 (4.87) | 18.44 (2.17) | 18.67 (2.85) |

Note: Volumes are presented in millilitres and left and right combined. \*Significantly reduced vs controls, <sup>a</sup>significantly reduced vs CBS, <sup>b</sup>significantly reduced vs nfVPPA, <sup>c</sup>significantly reduced vs PSP.

Partial correlations were conducted across the three patient groups to examine relationships between subcortical volumes and MLSE total score whilst controlling for estimated total intracranial volume. There was a significant negative partial correlation between pallidum volume and MLSE total score (*r*(35) = −.336, *N* = 38, *p*=.042) and a significant positive partial correlation between amygdala volume and MLSE total score (*r*(35) = .388, *N* = 38, *p*=.018).

We assessed the multivariate relationship between MLSE domain scores and structural scores (cortical thickness and subcortical volumes) of 83 nodes across the brain using canonical correlation analysis. We identified one significant pair of latent variables (MLSE-LV and Structure-LV, *r* = 0.5731 *p*<.001), see Figure 4. The Structure-LV expressed the highest loadings in the superior temporal cortex, prefrontal, inferior frontal and precentral regions, and in volumes of amygdala, hippocampus, putamen and caudate, with a tendency for left lateralisation. The MLSE-LV expressed all domains, with the highest loadings on motor speech, phonology and syntax domains, followed by semantic and auditory-verbal working memory domains (Figure 4A). The positive loading values indicated that higher performance on MLSE domains (more so motor speech, phonology and syntax domains) is associated with greater cortical thickness in the frontotemporal regions shown in Figure 4B and in volumes of amygdala, hippocampus, putamen and caudate (Figure 4C), with a tendency for left lateralisation.

**Figure 4.**
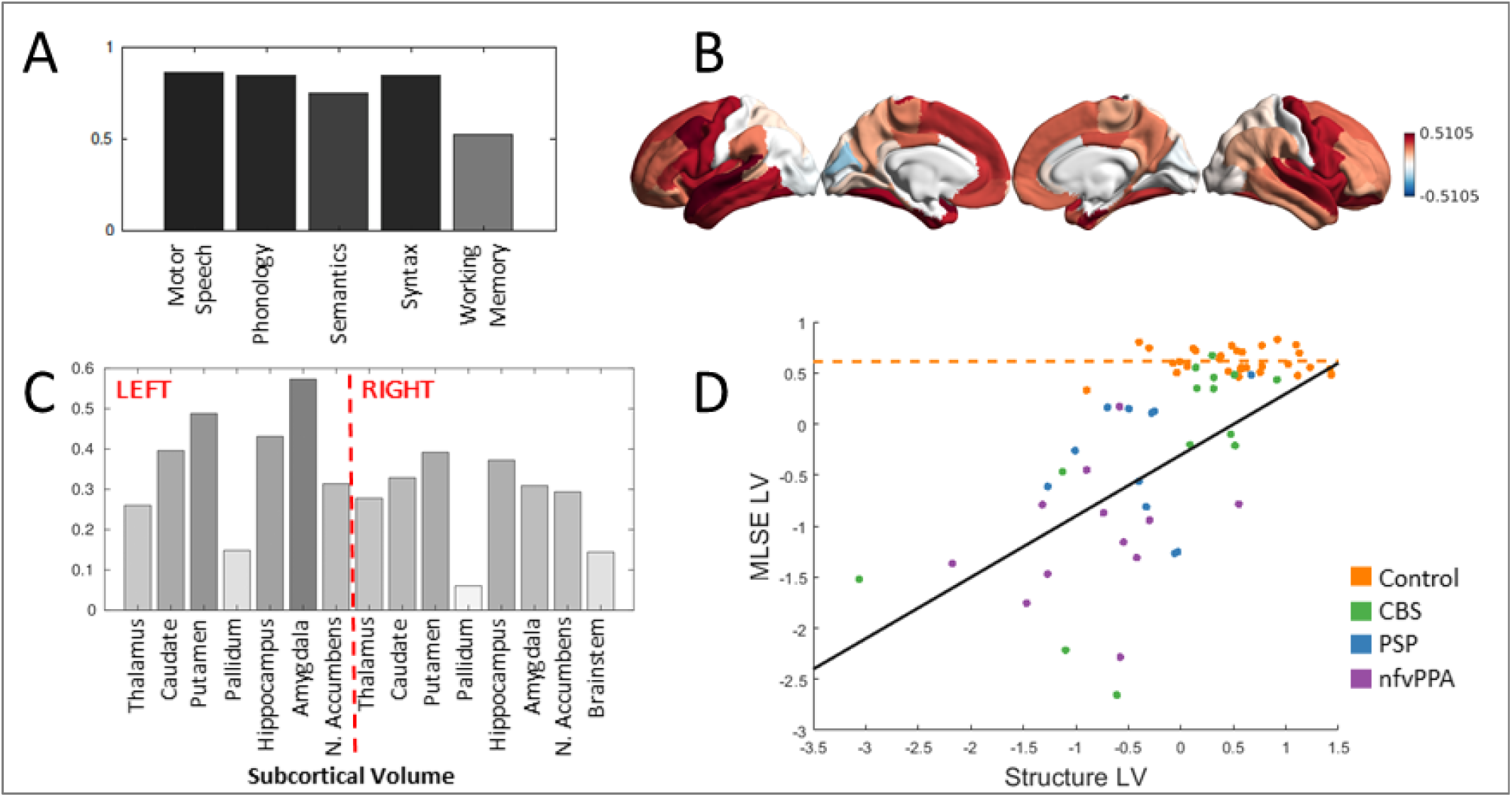
(A) The MLSE latent variable (MLSE-LV) loadings. The MLSE-LV expressed all domains, with the highest loadings on motor speech, phonology and syntax, followed by semantic and auditory-verbal working memory domains. (B) and (C) The Structure latent variable (Structure-LV) loadings. The Structure-LV expressed high loadings in the superior temporal cortex, prefrontal, inferior frontal and precentral regions and in volumes of amygdala, hippocampus, putamen and caudate, with a tendency for left lateralisation. (D) Visualisation of the interaction effect and the relationship between the MLSE-LV and the Structure-LV in the patients and controls. The relationship between MLSE-LV and Structure-LV was stronger in the patients relative to controls as visualised by black and orange trendlines, respectively, and confirmed formally by a significant interaction term (*r* = 0.562, *p*=.006). CBS, PSP and nfvPPA groups are colour coded.

To test whether the observed relationship between MLSE-LV and Structure-LV is differentially expressed between controls and patients, we constructed a second-level regression model with robust error estimates by including Structure-LV subject scores, group information and their interaction term as independent variables and MLSE-LV as dependent variable in addition to covariates of no interest (Figure 4D). We found evidence for a significant interaction (*r* = 0.562, *p*=.006) indicating a stronger relationship between MLSE-LV and Structure-LV in the patients relative to controls.

## Discussion

This study reveals the severity and structural correlates of language impairment in PSP and CBS, using the Mini Linguistic State Examination (MLSE). The PSP and CBS patients showed impaired performance on the MLSE domains of motor speech, phonological structure, semantic knowledge, and syntactic knowledge, but not auditory-verbal working memory. This pattern is similar to nfvPPA, and previous reports of CBS-NAV or PSP-SL (Burrell et al., 2018; Catricalà et al., 2019; Dodich et al., 2019). PSP and CBS were similar to each other in severity and range of language impairment.

We identified similar brain structural correlates of MLSE performance in PSP, CBS and nfvPPA. Multivariate analysis confirmed the association between a language component based on the MLSE domain scores (motor speech, phonology, and syntax loading most strongly) and a structural component (left-lateralised fronto-temporal cortical thinning and subcortical atrophy). Further, we found cortical thinning common to all three patient groups in pre-frontal and precentral gyri. This accords with previous research showing that motor speech, phonology, and syntactic ability are the most affected linguistic domains in PSP, CBS, and nfvPPA (Burrell et al., 2018; Dodich et al., 2019; Peterson et al., 2019), and supports the sensitivity of MLSE to structural changes associated with language effects of PSP and CBS.

This atrophy is consistent with findings of hypometabolism in the left inferior frontal gyrus in patients with PSP-SL and CBS-NAV (Dodich et al., 2019), implicating this region in the emergence of an nfvPPA-type language profile. Despite this consistency, there is considerable heterogeneity in patterns of structural and functional impairment in these disorders. For example, some CBS patients show a pattern of language impairment resembling the logopenic variant of PPA, with impaired complex sentence repetition (Dodich et al., 2019), together with bilateral parietal hypometabolism, possibly reflecting underlying Alzheimer’s disease pathology rather than corticobasal degeneration as the cause of CBS. Detailed analysis of linguistic impairment at the individual level in conjunction with pathological classification in PSP and CBS might provide more insight into the clinical-anatomical correlates of language impairment in these disorders.

The MLSE average assessment time was less than 20 minutes, but this was sufficient to confirm mild to moderate impairment in motor speech, semantic knowledge, phonological abilities, and syntactic ability in PSP and CBS (Burrell et al., 2018; Peterson et al., 2019). This confirms the MLSE as a quick language screening tool for patients with mixed cognitive and movement disorders. Many language tests assume good visual and motor functions (e.g. tasks featuring visual stimuli or which require writing). Such tasks might disadvantage PSP patients due to their oculomotor abnormalities or disadvantage both PSP and CBS patients due to motor deficits. This complicates interpretation of results from investigations of language in these disorders because scoring is often binary, the reasons for task failures can be unclear and can differ across disorders (Peterson et al., 2019; Picillo et al., 2019). The MLSE addresses this longstanding issue by incorporating an error-based scoring system to capture language-specific contributions to impaired test performance, enabling one to tease apart linguistic deficits from one another and from other impairments.

There are limitations to the present study. We do not have pathological validation in our sample although clinicopathological correlations of PSP are very high (>90%) (Gazzina et al., 2019). We have not examined phenotypic variance due to the small group sizes and insufficient power but note that our groups represent diverse phenotypes. The study did not aim to dissect phenotype specific patterns of atrophy of linguistic impairment, but rather exploit cohort variance to examine structure-function relationships. We recognise a possible selection bias for the patients scanned, with more severely impaired patients less likely to have undergone MRI. Thus, our imaging results may be more reflective of early-to-mid stage PSP/CBS/nfvPPA. Finally, the cross-sectional nature of this study precludes discussion of the progression of language profiles.

In conclusion, we find evidence for mild to moderate speech and language deficits in PSP and CBS which are similar in profile to nfvPPA. We have identified a shared anatomical substrate that correlates with linguistic impairment across these disorders, sensitive to MLSE profiling, consistent with the overlapping clinical and pathological spectrum of PSP, CBS, and nfvPPA.

## Data Availability

We invite requests for the anonymised data for academic (non-commercialised) purposes, to the corresponding author. Data sharing may be subject to restrictions on some data types to protect confidentiality

## Acknowledgements

The researchers thank the NIHR Join Dementia Research service for support provided during recruitment.

